# Use of a Continuous Single Lead Electrocardiogram Analytic to Predict Patient Deterioration Requiring Rapid Response Team Activation

**DOI:** 10.1101/2024.02.09.24302599

**Authors:** Sooin Lee, Bryce Benson, Ashwin Belle, Richard P. Medlin, David Jerkins, Foster Goss, Ashish K. Khanna, Michael A. DeVita, Kevin R. Ward

## Abstract

Identifying the onset of patient deterioration is challenging despite the potential to respond to patients earlier with better vital sign monitoring and rapid response team (RRT) activation. In this study an ECG based software as a medical device, the Analytic for Hemodynamic Instability Predictive Index (AHI-PI), was compared to the vital signs of heart rate, blood pressure, and respiratory rate, evaluating how early it indicated risk before an RRT activation. A higher proportion of the events had risk indication by AHI-PI (92.71%) than by vital signs (41.67%). AHI-PI indicated risk early, with an average of over a day before RRT events. In events whose risks were indicated by both AHI-PI and vital signs, AHI-PI demonstrated earlier recognition of deterioration compared to vital signs. A case-control study showed that situations requiring RRTs were more likely to have AHI-PI risk indication than those that did not. The study derived several insights in support of AHI-PI’s efficacy as a clinical decision support system. The findings demonstrated AHI-PI’s potential to serve as a reliable predictor of future RRT events. It could potentially help clinicians recognize early clinical deterioration and respond to those unnoticed by vital signs, thereby helping clinicians improve clinical outcomes.

**Author Summary:** Recognizing patient deterioration remains challenging even for experienced clinicians and nurses. RRTs can help mobilize resources to respond to patients earlier. However, determining when to activate RRTs is difficult. We retrospectively evaluated a software as a medical device, AHI-PI, compared the vital signs of heart rate, blood pressure, and respiratory rate to understand if AHI-PI could provide an earlier indicator of patient deterioration than vital signs. Our findings demonstrated AHI-PI’s potential to serve as a reliable predictor of future RRT events, before vital sign changes occur. This could potentially help clinicians recognize patients at risk for clinical deterioration and improve clinical outcomes through early targeted therapy or interventions.

## Introduction

Recognizing signs of early physiologic instability in hospitalized patients is critical to improving outcomes and preventing mortality. Failing to do so can contribute to unexpected transfers to a higher level of care, increased lengths of stay, and even unexpected deaths. Studies show that deliberate and consistent monitoring of vital signs improves early detection and clinical action.^1, 2^ However, measuring and documenting vital signs remain inconsistent and error prone in practice.^3, 4^ As a result, there may be delays in both recognizing and treating a patient at risk for deterioration. There are additional compounding factors that increasingly contribute to this problem. A few examples of such factors are nursing shortages, growing clinical workloads, patient comorbidities, data limitations, and resource constraints.^5^

As a mitigation strategy, hospitals in many countries and across the United States have developed rapid response systems designed to detect deterioration early and rapidly respond to those patients believed to be deteriorating. These systems have teams that are called by different names and include rapid response teams (RRTs), medical emergency response teams, high acuity response teams, code teams, etc.^6^ These response teams are typically multidisciplinary including physicians, nurses, respiratory therapists, and pharmacists supporting deteriorating patients to prevent potentially avoidable adverse events such as cardiopulmonary arrest.^7^ However, even with widespread adoption of RRTs, their effectiveness continues to be debated and evaluated.^6, 8-10^

The detection and activation of rapid response is prone to delays. A common cause of delay is caused by infrequent vital signs checks.^11^ Unexpected ICU admissions burden health care resources and are associated with substantial increases in cost of care, length of hospital stay and a nearly 20% increase in mortality.^12^ While there is potential for responding to patients earlier with better vital sign monitoring and RRTs, identifying the onset of patient deterioration is challenging. This naturally makes determining when to activate an RRT difficult. Clinical decision support systems that help clinicians recognize clinical deterioration prior to changes in traditional vital signs could be valuable for early intervention. Desirable characteristics of such a system would include easy automation, continuous output, simple interpretation, and no significant change or burden added to clinical workflow.

In this study, we evaluate the Analytic for Hemodynamic Instability-Predictive Index (AHI-PI: Fifth Eye, Ann Arbor, MI), a FDA approved software as a medical device (SaMD) that uses a continuous ECG waveform and extracted heart rate variability (HRV) to predict hemodynamic instability.^13-15^ Changes in HRV have been demonstrated to reflect changes in the autonomic nervous system in the setting of many states of critical illness and injury including hemorrhage, sepsis, cardiogenic shock, respiratory failure, and others with these changes occurring prior to overt decompensation.^16-23^ In previous studies, we demonstrated its ability to predict hemodynamic instability (a combination of tachycardia and hypotension) with high sensitivity and specificity with average lead times greater than 3 hours.^15, 24^

AHI-PI automates the extraction and analysis of ECG patterns to reflect the compensatory burden on the autonomic nervous system to provide information regarding the patient’s predicted future risk for clinical deterioration based on the known physiologic relationship of HRV, the autonomic nervous system, and the cardiovascular system.^16, 19^ It also includes a signal quality assessment and processing of extracted patterns through a pretrained classification model that embeds nonlinear HRV complexity and ECG morphologic features into a single output.^13-15^

AHI-PI builds on this output and updates every two minutes, producing one of three types of outputs, red, yellow, or green, indicating high, moderate, or low risk respectively of a future episode of hemodynamic instability.^24^ Hemodynamic instability is defined as a combination of heart rate greater than or equal to 100 beats per minute and a systolic blood pressure (SBP) less than 90 mmHg or mean arterial pressure (MAP) of less than 70 mmHg.

We examine and assess the ability of AHI-PI in predicting the need for an RRT prior to its activation by comparing it to changes in the vital signs of heart rate and blood pressure, but also now add respiratory rate for hospitalized patients who were undergoing continuous ECG monitoring and in which an RRT was activated.

## Methods

This was a retrospective single-center observational cohort study conducted at the University of Michigan, a quaternary academic health system in Ann Arbor, Michigan, between August 2019 and April 2020. The study dataset included consecutive hospitalized adult (≥ 18 years) patients who were undergoing continuous ECG monitoring on telemetry, stepdown and intensive care units and for whom an RRT was activated.

The study was approved by the University of Michigan Institutional Review Board (HUM00092309). Due to its retrospective design and use of deidentified data, a waiver of consent was granted.

Patient data including demographic and vital signs were extracted from Epic Systems (Verona, WI) electronic health record (EHR). ECG waveform data were obtained from an external data warehouse (EDW) that collects, stores, and maintains a high-resolution physiologic signal database of patients including real-time physiologic signals and waveforms such as ECG. The ECG was sampled at 240 Hz for this study.

### RRT event

The study defined an RRT event as a rapid response event including cardiac arrest (code calls) as recorded in the EHR. AHI-PI outputs and EHR-recorded vital signs within 48 hours before each RRT event were analyzed in this study to account for out-of-range vital signs that were documented before activation of the RRT.^25-27^ To ensure that no redundant AHI-PI or vital signs data were used for patients having more than one RRT event, only those at least 48 hours apart were chosen, starting with the first one in chronological order.

### Study dataset

The study data initially contained a total of 168 RRT events which were at least 48 hours apart. Patients with any exclusions (i.e., heart transplants, ventricular assist device, and chronic arrhythmias such as atrial fibrillation) were removed from the analysis as these conditions makes HRV an unreliable predictor.^28, 29^ Patients without any valid AHI-PI outputs in the previous 48 hours prior to the event were also excluded from the analysis leaving 110 RRT events for analysis. Blinded to AHI-PI’s results, two physicians from Michigan Medicine then reviewed each event to determine if each of the RRT calls resulted in a true escalation of care (e.g., need for transfer to the ICU, institution of vasopressors, mechanical or noninvasive ventilation for respiratory failure, blood transfusion, or cardiopulmonary resuscitation at the time as recorded in the EHR). After clinician review, there were 96 RRT events from 91 patients identified (Fig 1).

**Fig 1.**
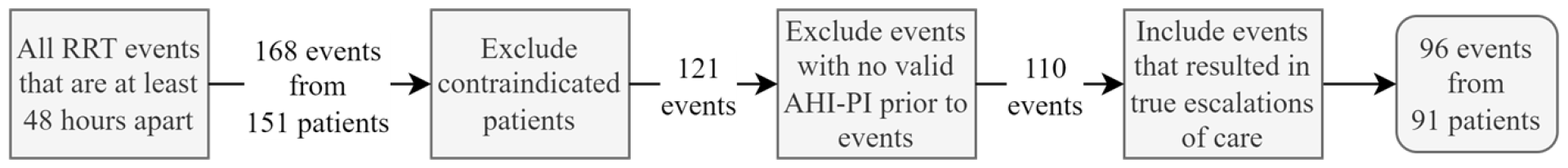
Identifying the study cohort and dataset.

### AHI-PI and vital signs risk indication

The study defined the presence of risk indication as having AHI-PI high or moderate risk output within 48 hours before a given RRT event. That is, if AHI-PI had one or more high or moderate risk outputs within 48 hours before an event, then AHI-PI was said to have indicated risk before the event. For vital signs, the presence of risk indication was conservatively defined as having hypotension (SBP < 90mmHg or MAP < 70mmHg) combined with either tachycardia or tachypnea (pulse ≥ 100 beats per minute or respiratory rate > 25 breaths per minute). That is, if hypotension and tachycardia or hypotension and tachypnea were recorded within 48 hours before the event, then vital signs were said to have indicated risk. The combination of tachycardia or tachypnea combined with hypotension, in our opinion, gives high confidence that the “out-of-range” period represents a conservative ground truth indicator of the presence of hemodynamic instability.^30, 31^

### AHI-PI and vital signs initial risk indication time

In addition to looking at the presence of risk indication in AHI-PI and vital signs in the previous 48 hours of an RRT event, the study calculated how long before the event AHI-PI and vital signs first indicated risk within the analysis timeframe (Fig 2). For AHI-PI, the initial risk indication time was measured as the distance from the RRT event time to the first time AHI-PI produced high or moderate risk output within 48 hours before the event. For vital signs, the initial risk indication time was measured as the distance from the event time to the first time both hypotension and tachycardia or tachypnea were present within the previous 48 hours of the event. For example, if tachycardia was recorded 10 minutes after presence of hypotension in the EHR, then the time when tachycardia was recorded would be used to calculate vital signs’ initial risk indication time.

**Fig 2.**
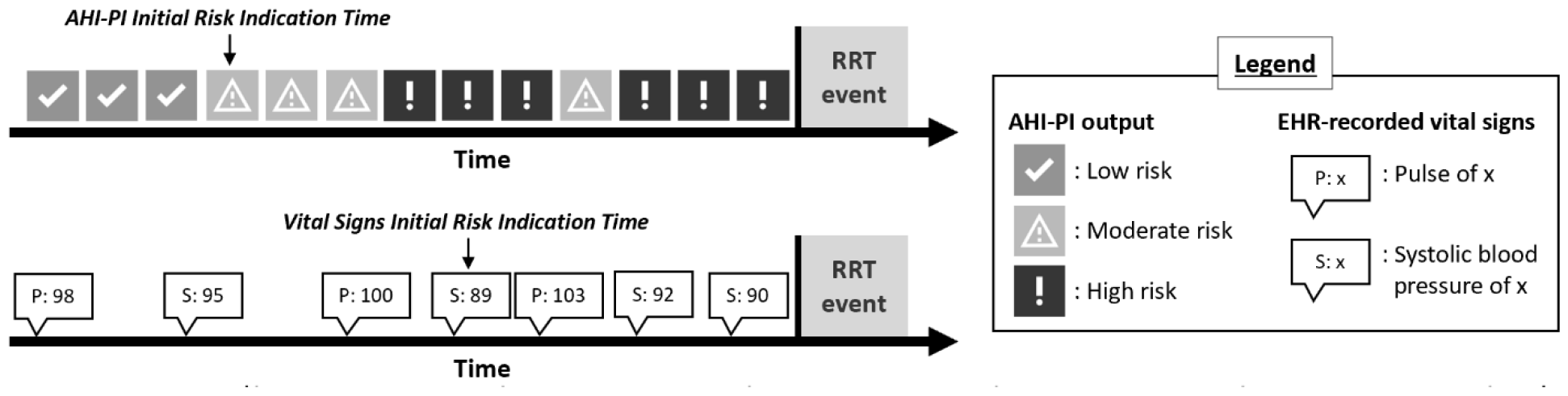
AHI-PI and vital signs initial risk indication time. This example shows hypotension and tachycardia having occurred prior to RRT activation.

### Case-control study

Even though the main goal of the study was to evaluate AHI-PI in situations that required RRTs specifically, the study also sought to evaluate AHI-PI’s performance in patients where RRTs were not called by examining a case-control cohort. For each of the patient-encounters included in this RRT study, a cohort without any RRT event was identified and paired with it for comparison. Each control case was matched 1:1 to each RRT cases by age (+/- <6 years), sex, unit type, primary diagnosis, and ECG monitoring duration. Matching for the ECG monitoring duration allowed assigning each control case an event time such that the event happened proportionately at the same time as its matched RRT case. A more detailed description of the steps taken to identify control cases is in Appendix A.

A two-sample t-test and conditional logistic regression were used to compare percentages of AHI-PI risk indication within 48 hours before the events in the control and RRT cases. In all cases, AHI-PI was running and executed silently in a background environment not accessible to clinicians or care teams; therefore, no interventions to mitigate risk identified by AHI-PI were taken at any stage.

## Results

The mean patient age was 61.25 ± 14.36 years. Patient demographics are provided in Table 1. AHI-PI indicated risk in 92.71% (89) of the 96 RRT events (Fig 3). All 89 events (subgroup 1) had AHI-PI high or moderate risk outputs within 48 hours before the RRT. In contrast, EHR documented vital signs indicated risk in 41.67% (40) of the 96 RRT events, with all 40 events having documented hypotension and tachycardia or tachypnea in the previous 48 hours. In these 40 events, both vital signs and AHI-PI indicated risk (subgroup 1A). Where documented vital signs indicated no risk before the RRT event, AHI-PI indicated risk in 51.04% (49) of events (subgroup 1B). The study found 2.08% (2) of the 96 RRT events showed risk indication with vital signs’ and without AHI-PI’s (subgroup 2A) and 5.21% (5) of the 96 events had neither vital signs nor AHI-PI presented risk indication prior to the events (subgroup 2B). Results are summarized in the contingency table in Figure 3 and reasons for RRT calls are shown in Table 2.

**Table 1.** Study cohort demographics.

| Total Number of Patients |  | 91 |
| --- | --- | --- |
| **Sex** | *Male (%)* | 43 (47.25%) |
|  | *Female (%)* | 48 (52.75%) |
| **Age (years)** | *Mean (Standard Deviation (SD))* | 61.25 (14.36) |
|  | *Range* | 26.39, 89.63 |
| **Race** | *White* | 67 |
|  | *Black or African American* | 15 |
|  | *Asian* | 3 |
|  | *American Indian or Alaska Native* | 2 |
|  | *Native Hawaiian and Other Pacific Islander* | 1 |
|  | *Other* | 2 |
|  | *Unknown or Not Reported* | 1 |
| **Ethnicity** | *Non-Hispanic* | 87 |
|  | *Hispanic* | 3 |
|  | *Unknown or Not Reported* | 1 |

**Table 2.** Summary of reasons for the RRT events.

|  | Subgroup 1A | Subgroup 1B | Subgroup 2A | Subgroup 2B | All RRT Events |
| --- | --- | --- | --- | --- | --- |
| **Respiratory distress/failure related** | 12 | 19 | 1 | 0 | 32 |
| **Hypotension related** | 17 | 8 | 1 | 1 | 27 |
| **Respiratory distress/failure and hypotension related** | 2 | 1 | 0 | 1 | 4 |
| **Dysrhythmia related** | 2 | 7 | 0 | 0 | 9 |
| **Hypotension and dysrhythmia related** | 1 | 0 | 0 | 0 | 1 |
| **Cardiac arrest related** | 5 | 9 | 0 | 2 | 16 |
| **Hemorrhage related** | 1 | 0 | 0 | 1 | 2 |
| **Stroke related** | 0 | 2 | 0 | 0 | 2 |
| **Seizure related** | 0 | 2 | 0 | 0 | 2 |
| **Chest pain related** | 0 | 1 | 0 | 0 | 1 |
| **Total** | 40 | 49 | 2 | 5 | 96 |

**Fig 3.**
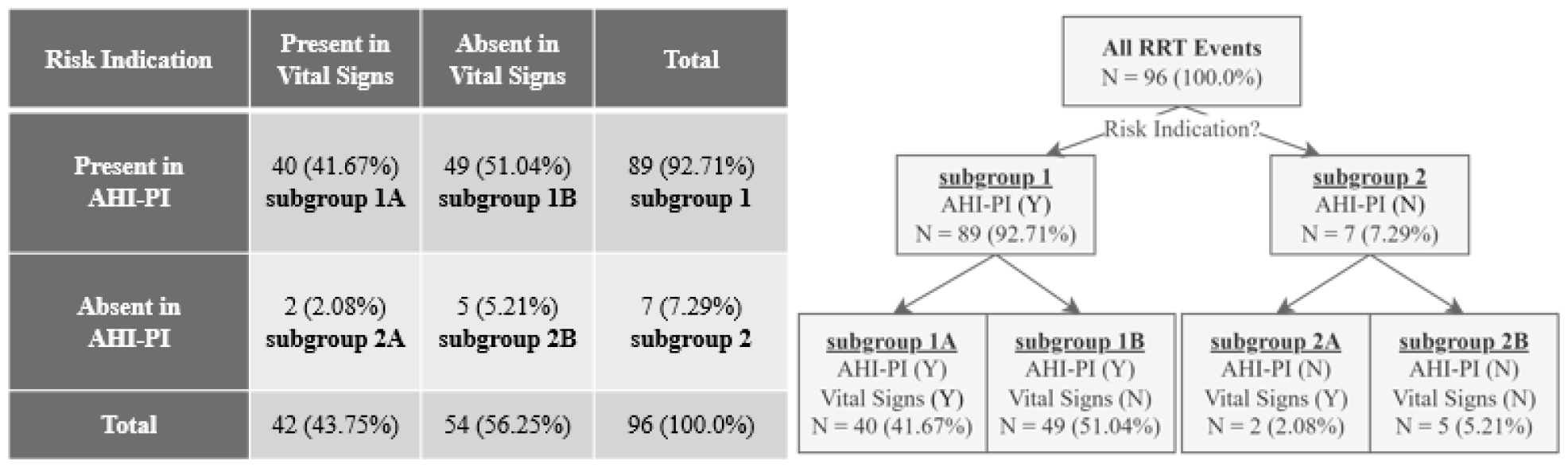
Table and flowchart summarizing presence (Y) / absence (N) of risk indication.

The exact analysis timeframes varied across the 96 RRT events because they depended on the durations of ECG monitoring preceding the events. The median analysis timeframe, or ECG monitoring duration, across the events was 45.88 hours, with a mean of 30.38 hours. Overall, AHI-PI outputs were available and valid the majority of time, with a median percentage of available outputs being 89.29%. That is, AHI-PI produced few unavailable, or invalid, outputs across the events during the analysis timeframes. The distributions of ECG monitoring durations as well as of the counts and percentages of available AHI-PI outputs for the 96 events are shown in Table 3.

**Table 3.** Numerical summaries of ECG monitoring durations and of the counts and percentages of available AHI-PI outputs.

|  | **ECG Monitoring Duration (in hours)** | **Count of Available AHI-PI Outputs** | **% Available AHI-PI Outputs** |
| --- | --- | --- | --- |
| **Mean (Standard Deviation)** | 30.38 (19.91) | 727.86 (539.10) | 80.18 (22.73) |
| **Median** | 45.88 | 750.50 | 89.29 |
| **Interquartile Range** | 7.92, 48.0 | 140.0, 1,293.25 | 72.82, 96.32 |

### AHI-PI initial risk indication time by subgroup

A summary of AHI-PI initial risk indication times for subgroups 1, 1A, and 1B can be found in Figure 4. Across all 89 events (subgroup 1), AHI-PI’s initial risk indication time was 26.48 hours before the events, on average (median: 28.57 hours). The mean initial risk indication times were 30.54 and 23.17 hours for subgroups 1A and 1B, respectively (medians: 42.67 and 21.80 hours). Comparing subgroup 1A and subgroup 1B using Mann-Whitney U test, the study did not find sufficient evidence to conclude that the initial risk indication time for AHI-PI in subgroup 1A is different from that in subgroup 1B (p-value: 0.1113).

**Fig 4.**
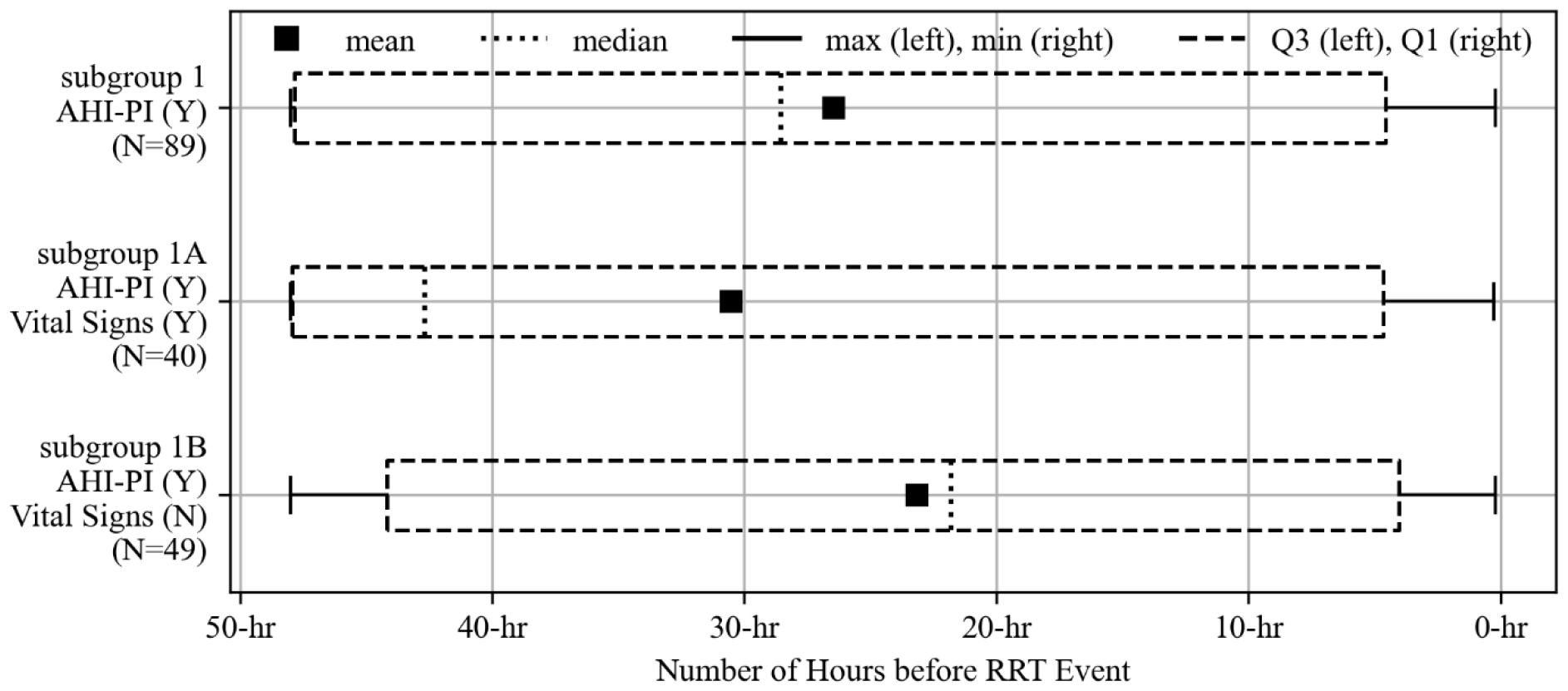
Distributions of AHI-PI initial risk indication times for the three subgroups

### AHI-PI risk indication in and outside of ICU

Of the 96 RRT events, 60 (62.5%) occurred within ICUs and 36 (37.5%) outside of ICUs. The study examined the 60 and 36 events, separately, and found their results to be consistent with the results from all care units regarding how AHI-PI and vital signs indicated risk. A higher proportion of events had risk indication by AHI-PI than by vital signs in and outside of ICUs.

Unit specific distributions of AHI-PI’s initial risk indication times are shown in Figure 5 and exhibit similar distributions as in Figure 4. Across all of subgroup 1, the mean AHI-PI initial risk indication times were 28.00 and 24.25 hours before the RRT events for the events in and outside of ICUs, respectively. For subgroup 1A, the mean initial risk indication times were 30.05 and 31.68 hours in and outside of ICUs. The mean times were 25.71 and 20.53 hours in and outside of ICUs for subgroup 1B, respectively.

**Fig 5.**
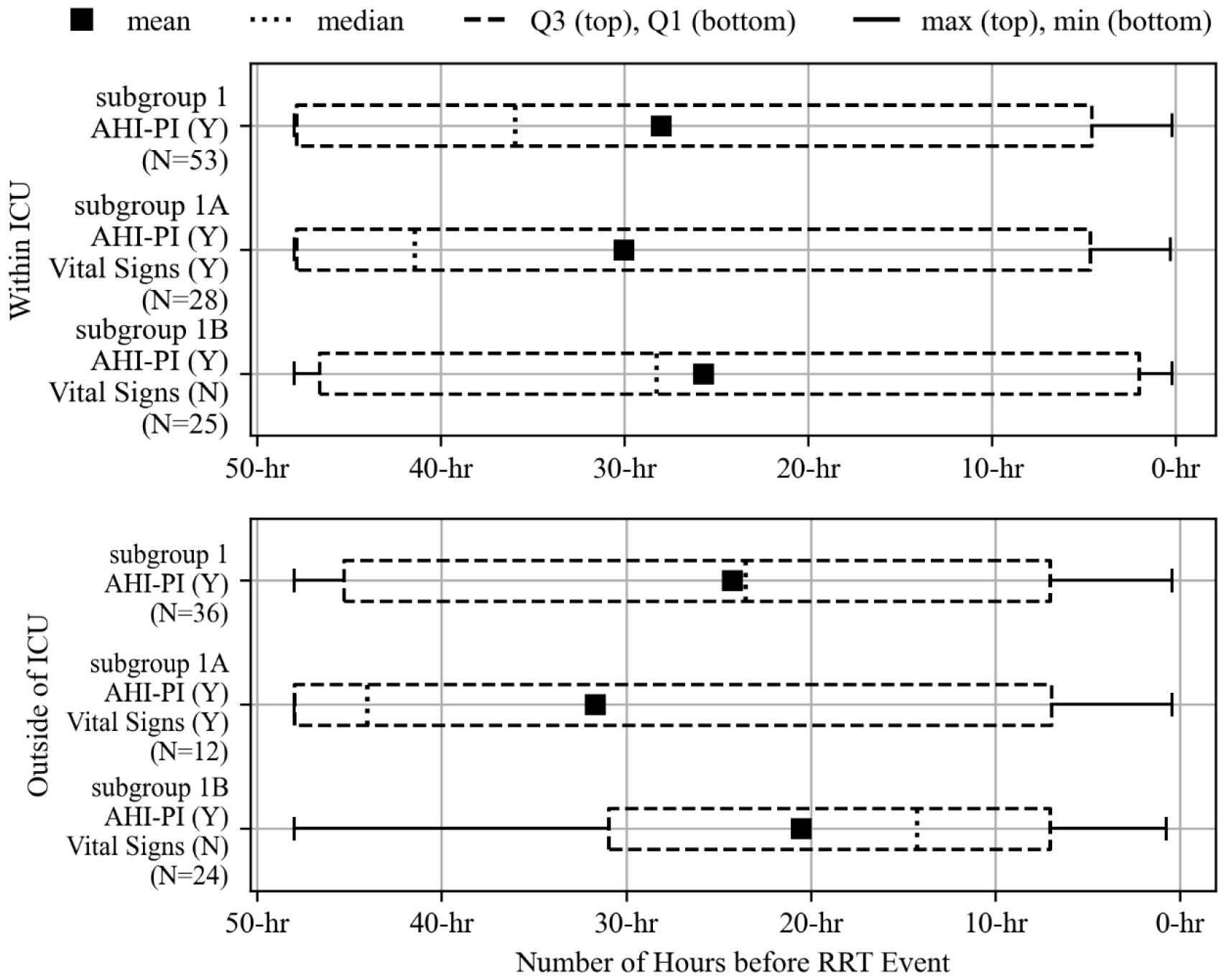
Distributions of AHI-PI initial risk indication times for the three subgroups by unit type

### Comparison between AHI-PI and vital signs for subgroup 1A

As previously described and shown in Figure 3, there were 40 RRT events where both AHI-PI and vital signs indicated risk (subgroup 1A). These 40 events’ initial risk indication times distributions for AHI-PI and vital signs are shown in Figure 6. AHI-PI’s initial risk indication time was 30.54 hours, on average (median: 42.67 hours, range: [0.26 hour, 48.0 hours], interquartile range: [4.66 hours, 47.95 hours]). In contrast, the mean vital signs initial risk indication times was 19.26 hours before the events, respectively (median: 10.87 hours, range: [0.09 hour, 47.92 hours], interquartile range: [1.51 hours, 40.18 hours]). The study found sufficient evidence that AHI-PI’s initial risk indication time was earlier than vital signs’ (p-value: 0.0062 (Two-sample t-test)) and that their distributions are not equal (p-value: 0.0022 (Mann-Whitney U test)). Additionally, in 32 of the 40 events, AHI-PI indicated risk before it was evident in vital signs. Comparing these 32 events’ initial risk indication times, AHI-PI first indicated risk 15.87 hours before vital signs, on average (median: 8.20 hours).

**Fig 6.**
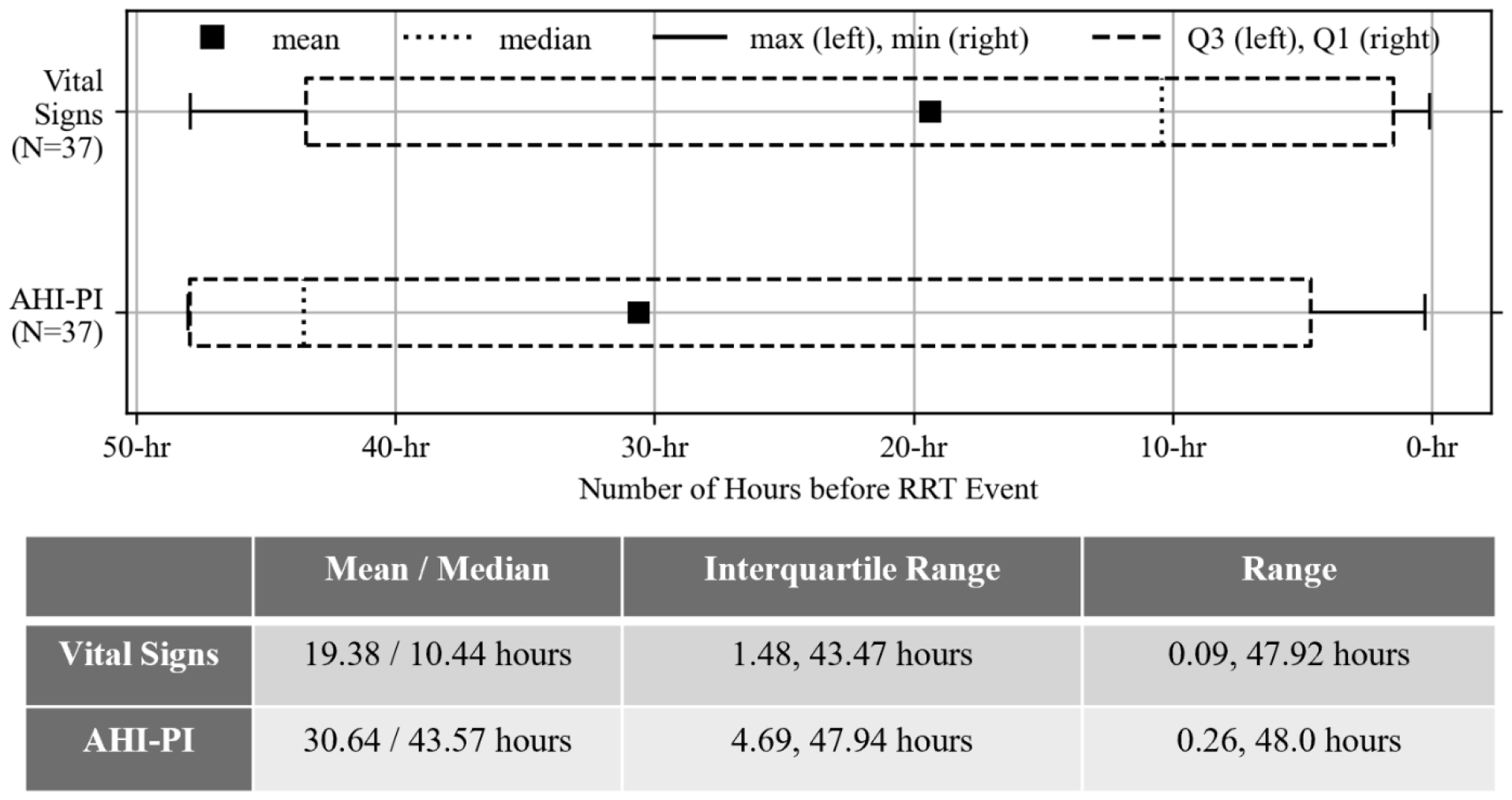
Boxplots showing the distribution of vital signs initial risk indication times (top) as well as the distribution of AHI-PI initial risk indication times (bottom) for subgroup 1A.

The study also found that, once AHI-PI indicated risk for the first time, its outputs often showed high or moderate risk leading up to the RRT events. As shown on the far right of Figure 7, the mean and median percentage of risk outputs between the initial risk indication time and event time were 67.57% and 76.36%, respectively (interquartile range: [40.21%, 100.0%]). The study also looked at the distributions of SBP, MAP, respiratory rate, and heart rate values after vital signs’ initial risk indication time, leading up to the event time (Figure 7). The mean and median SBP were 108 mmHg and 105 mmHg, respectively. The mean MAP was 77 mmHg (median: 75 mmHg). The mean and median respiratory rates were 24 and 22 breaths per minute. The mean pulse was 101 bpm, with a median of 101 bpm.

**Fig 7.**
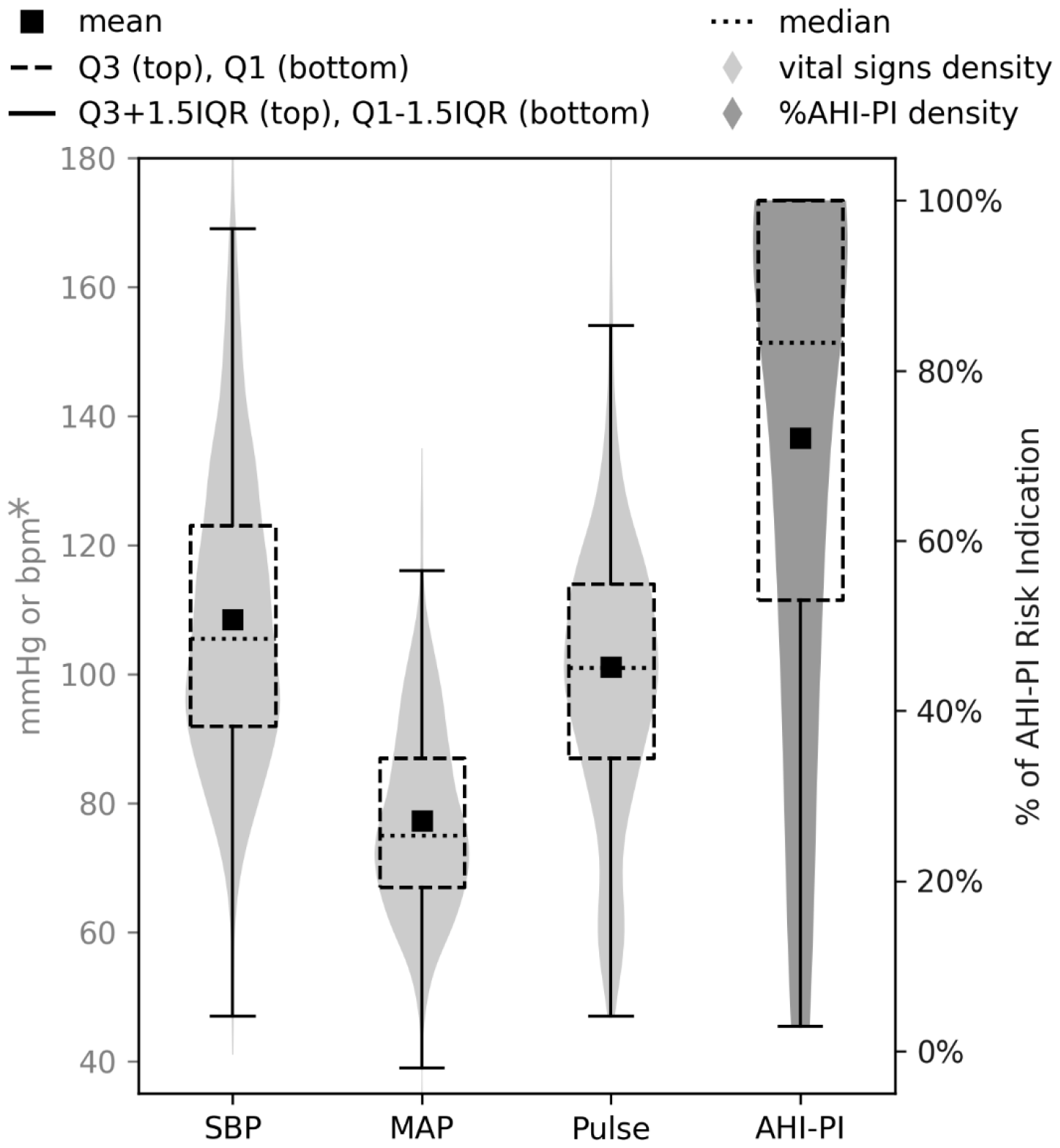
Distributions of EHR-recorded SBP, MAP, respiratory rate, and pulse values between the vital signs initial risk indication time and event time and of percentages of AHI-PI risk indications between AHI-PI initial risk indication time and event time (subgroup 1A). Y-axis label ‘bpm*’ is either beats per minute (heart rate) or breaths per minute (respiratory rate) depending on the context.

In addition to finding that AHI-PI indicated risk earlier than vital signs when all units are combined (Figure 6), the study also found this within ICUs and outside of ICUs. This consistent finding is depicted in Figure 8. In fact, for subgroup 1A where both AHI-PI and vital sign changes occurred before RRT, AHI-PI was earlier than vital signs in 23 of the 28 events within ICUs by an average of 13.77 hours and in 9 of the 12 events outside of ICUs by an average of 21.25 hours.

**Fig 8.**
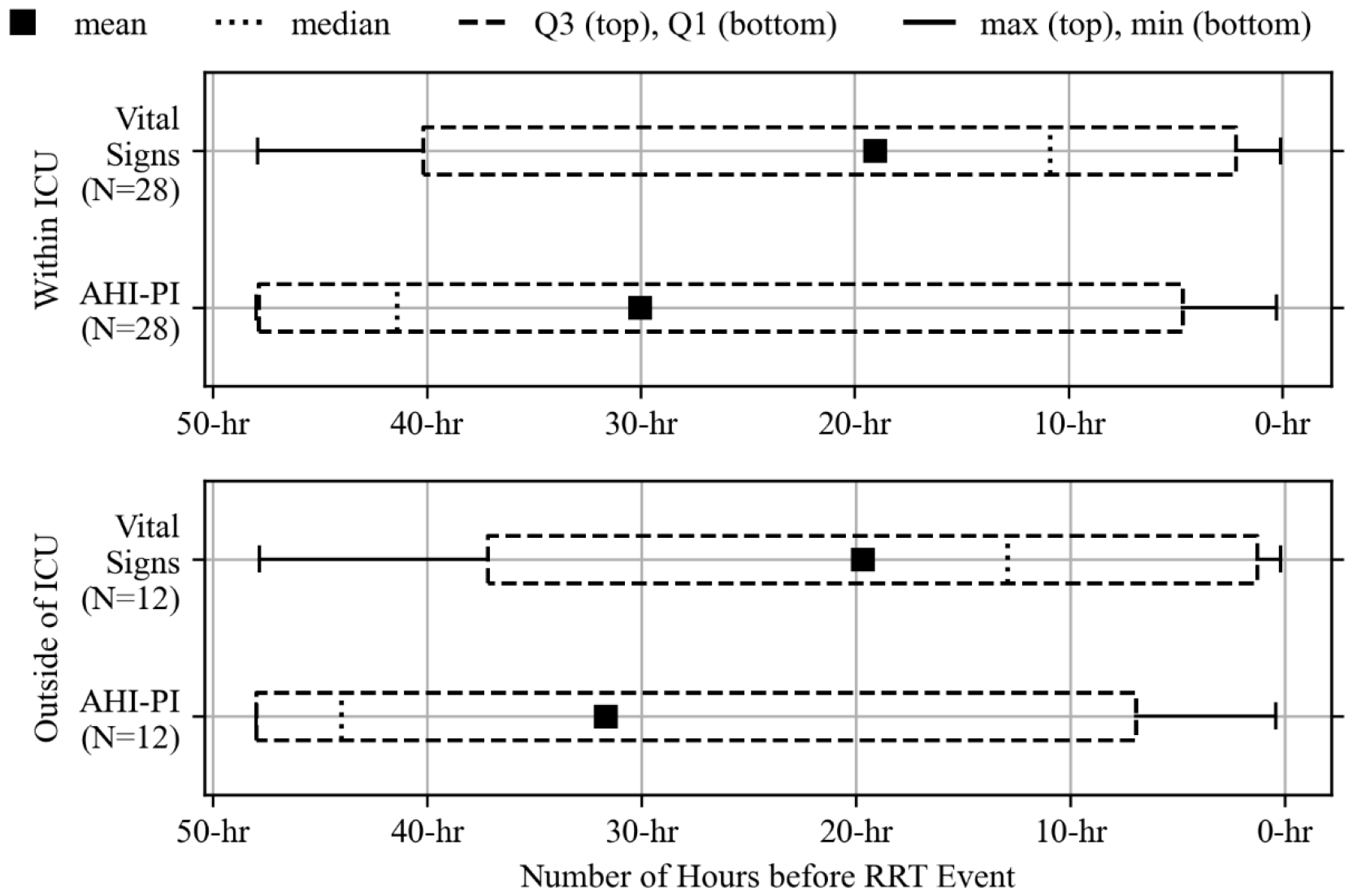
Distributions of initial risk indication times for vital signs and AHI-PI by unit type (subgroup 1A).

### Case-control study

Lastly, the study compared the percentages of AHI-PI risk indication within 48 hours before the events in the RRT group to those in the control group by matching each control case 1:1 to each RRT case by age, sex, unit, diagnosis, and ECG monitoring duration. The study found that the matched control group of 96 cases had substantially fewer AHI-PI risk outputs, compared to the RRT group, overall. As shown in Table 4, the median percentage of risk indication was 12.60% for the control group, meaning that at least half of the controls had 12.60% or less of their AHI-PI outputs indicating risk. On average, 33.12% of the outputs indicated risk across all the control cases. On the other hand, the median percentage of risk indication outputs was 48.85% for the RRT group, with a mean of 49.56%. The average percentage of AHI-PI risk indication for the control group is noticeably shifted upwards, compared to the RRT group whose average is close to its median. This is because matching each control case 1:1 to each of the RRT cases by characteristics such as unit type and primary diagnosis methodically led the control group to contain patients who were at similar risk for RRT as the actual RRT group. That is, while some of those in the control group did experience hemodynamic deterioration, none required RRTs.

**Table 4.** Numerical summaries of the percentages of AHI-PI risk indication for RRTs and for controls.

|  | **RRTs** | **Controls** |
| --- | --- | --- |
| **Mean (Standard Deviation)** | 49.56 (38.95) | 33.12 (38.65) |
| **Median** | 48.85 | 12.60 |
| **Interquartile Range** | 10.38, 97.28 | 0.0, 71.71 |

By design, what mainly distinguishes the control group from the RRT group is that the controls did not have RRT events. In fact, 65 cases in the control group had at least one AHI-PI risk indication at some point before the assigned event times. Of the 65, 15 (23.08%) showed out-of-range vital signs before the event times, and 53 (81.54%) received fluids as a bolus or pressors during their encounters (but no RRT activation). A total of 9 out of the 65 did not receive such interventions and did not have out of range vital signs. Even so, the study found sufficient evidence that the percentage of AHI-PI risk indication was lower in the control group, compared to the RRT group, on average (p-value: 0.0019 (Two-sample t-test)).

The conditional logistic regression results are summarized in Table 5. The study found that the events requiring RRTs were more likely to have AHI-PI risk indication, compared to the events where RRTs were not needed or avoided. Specifically, compared to those without any AHI-PI risk indication, those with AHI-PI risk indication had 9 times the odds of having RRT events (odds ratio: 9.0).

**Table 5.** Comparison between RRTs and controls.

| **AHI-PI Risk Indication** | **RRTs N (%)** | **Controls N (%)** | **Odds Ratio (95% CI)** | **P-value** |
| --- | --- | --- | --- | --- |
| **Absent** | 7 (7.29) | 31 (32.29) | reference | - |
| **Present** | 89 (92.71) | 65 (67.71) | 9.0 (2.73-29.67) | 0.0003 |

## Discussion

Prevention of morbidity and mortality using RRTs can only work if high risk deteriorating patients are rapidly and reliably identified, and RRT calls are triggered. Few studies have worked on preventing physiologic decline before achieving the triggering criteria. While evidence exists that changes in vital signs can be helpful in the early identification of patients who may benefit from an RRT, identifying the early onset of clinical deterioration by vital sign measurements can be challenging due to various factors such as infrequent, inaccurate, or delayed vital signs documentation and suboptimal nursing to patient ratios.^5^ This can make it difficult to determine when to activate an RRT. As such there continues to be a need to develop clinical decision support tools that could optimize RRT utilization and efficacy by recognizing the potential for clinical deterioration at earlier time points. While largely theoretical, earlier identification of patients in need would allow for assessment and interventions that could potentially reduce the need for transfer to the ICU or increase earlier transfer to the ICU resulting in reduced total ICU stay and prevent catastrophic events such as cardiac arrest.

Before any prospective use of AHI-PI as a clinical decision support tool for RRT activation, the study sought to evaluate AHI-PI’s potential to contribute as such a support system using a retrospective analysis. The study examined patients in whom a RRT activation occurred (including code events for cardiac arrest), among all consecutive non-contraindicated adult patients with available ECG monitoring data. The study analyzed and compared AHI-PI to vital signs for the potential to serve as an earlier indicator and derived insights from these RRT events that have important implications in support of AHI-PI’s efficacy as a clinical decision support system.

Of the 96 RRT events included in the study, a significantly higher proportion of the events had risk indication by AHI-PI (92.71%; subgroup 1) than by vital signs (38.54%). All the events for which vital signs indicated risk in advance were also predicted by AHI-PI (subgroup 1A). Only 2 of all 96 RRTs (2.08%; subgroup 2A) were preceded by abnormal vital signs and not AHI-PI. Additionally, AHI-PI’s initial risk indication time was more than a day in advance of the events, on average. These findings support AHI-PI’s potential to provide additional clinical insights hours in advance of the need for RRT, which may help clinicians intervene ahead of time to avoid deterioration. Furthermore, all these findings were consistent among both events within ICUs and events outside of ICUs. The consistency across units supports AHI-PI’s potential as a clinical decision support system.

The study also found that AHI-PI indicated risk for the majority of the events without vital signs’ risk indication (subgroup 1B). This finding supports that AHI-PI could be used to help respond to patients who are unnoticed by vital signs. When looking specifically at the events with risk indication by both AHI-PI and vital signs (subgroup 1A), the study found that AHI-PI’s initial risk indication times were often earlier and more consistent in risk indication, compared to vital signs, after their respective initial indication times. These findings demonstrate AHI-PI’s potential to serve as a reliable predictor of future RRT events. Especially considering the continuous nature of AHI-PI, it could help clinicians recognize clinical deterioration sooner than when relying solely on vital signs, helping them improve clinical outcomes. Code and RRT responses from the ICUs with documentation of such events were included in this study. This is important given the increasing use of tele-ICU services and for ICUs which may be less resourced than academic medical centers.^32^

The types of RRT events (Table 2) are, for the most part, indicative of physiology that would impact and produce early changes in the autonomic nervous system and heart rate variability thus making AHI-PI a potentially useful clinical decision support tool. However, events such as cardiac arrest, certain sudden dysrhythmias, stroke, pulmonary embolism, anaphylactic reactions, and others where an RRT may be called may not lend themselves to prediction and early intervention based on HRV monitoring depending on the etiology and suddenness of the event as well as recognition of symptoms by providers. AHI-PI’s performance in some of these conditions were mixed.

A case-control study was conducted to understand and evaluate how AHI-PI performed in situations without RRT calls, compared to situations with them. In general, the control group had substantially less AHI-PI risk indicators, compared to the RRT group. This further supports that AHI-PI may reliably work as part of a clinical decision support system in detecting the need for an RRT or code team prior to activation.

AHI-PI is designed to be a clinical decision support tool, providing health care providers an advanced indicator when patients may require closer evaluation or re-evaluation. While statistically significant differences exist between AHI-PI and vital signs and AHI-PI indicated an increased need for an RRT (9 times greater risk) compared to controls, there is some overlap in performance between AHI-PI and vitals. However, as a continuous output variable, AHI-PI is expected to act as a powerful complement to vital signs and especially intermittent vital signs.

AHI-PI may also compliment EHR based early warning systems which in combination could increase accuracy, provide greater insight into the underlying cause of the alert, and assist in more rapid adjudication of the alarms.^33-35^ Furthermore, since AHI-PI is approved for use with various single lead wearable ECG patches, it may allow more intensive and informative monitoring of floor level patients who are monitored much less frequently.

## Limitations

The study used retrospective data, which were not collected or designed specifically for this study. This could mean that the data contained unknown biases or confounding factors that the study cannot account for. The study also did not take into account medications that were given to prevent deterioration in the control group. Additionally, although respiratory rate has been included in this analysis, its measurement through either bedside patient assessment by care providers or by bedside monitors using traditional thoracic impedance technology has demonstrated significant disagreement with true respiratory rates.^36-42^ Patients with heart transplants, ventricular assist device, and chronic persistent arrhythmias such as atrial fibrillation were excluded forming an additional limitation to this study as these are contraindicated for the AHI-PI analytic since they are known to limit or interfere with the determination of HRV.

Lastly, because only RRT events were evaluated, the sensitivity and specificity of AHI-PI cannot be determined. To what extent use of AHI-PI could have reduced RRT and code events by prompting increased vigilance and earlier interventions is unknown. However, the data presented supports the next required step which is a prospective study that examines AHI-PI’s efficacy in detecting the need for an RRT prior to activation will be needed.

## Conclusion

The study examined the ability of an ECG based SaMD product called AHI-PI as a clinical decision support system in detecting decompensation events requiring an RRT. The findings from this study demonstrated AHI-PI’s potential for identifying the need for RRTs early than changes in vital signs. Accordingly, AHI-PI could potentially help clinicians recognize early clinical deterioration, take necessary measures, and improve clinical outcomes. The data from this study supports future prospective trials.

## Data Availability

This data is owned exclusively by University of Michigan and was used for this analysis with an established data use agreement between Fifth Eye Inc. and the University of Michigan under a cleared IRB. Please reach out to the following University of Michigan personnel for your access to data. Phil Jacokes, Managing Director, Weil Institute

